# SWIFT: A Deep Learning Approach to Prediction of Hypoxemic Events in Critically-Ill Patients Using SpO_2_ Waveform Prediction

**DOI:** 10.1101/2021.02.25.21252234

**Authors:** Akshaya V. Annapragada, Joseph L. Greenstein, Sanjukta N. Bose, Bradford D. Winters, Sridevi V. Sarma, Raimond L. Winslow

## Abstract

Hypoxemia is a significant driver of mortality and poor clinical outcomes in conditions such as brain injury and cardiac arrest in critically ill patients, including COVID-19 patients. Given the host of negative clinical outcomes attributed to hypoxemia, identifying patients likely to experience hypoxemia would offer valuable opportunities for early and thus more effective intervention. We present SWIFT (SpO_2_ <u>W</u>aveform <u>I</u>CU <u>F</u>orecasting <u>T</u>echnique), a deep learning model that predicts blood oxygen saturation (SpO_2_) waveforms 5 and 30 minutes in the future using only prior SpO_2_ values as inputs. When tested on novel data, SWIFT predicts more than 80% and 60% of hypoxemic events in critically ill and COVID-19 patients, respectively. SWIFT also predicts SpO_2_ waveforms with average MSE below .0007. SWIFT provides information on both occurrence and magnitude of potential hypoxemic events 30 minutes in advance, allowing it to be used to inform clinical interventions, patient triaging, and optimal resource allocation. SWIFT may be used in clinical decision support systems to inform the management of critically ill patients during the COVID-19 pandemic and beyond.

## Introduction

Hypoxemia, or a decrease in blood oxygen saturation, is a common symptom in critically ill patients, with a multinational, multicenter study finding that hypoxemia is a significant risk factor for mortality, with prevalence greater than 50% in ICU patients (SRLF Trial Group, 2018). Severe hypoxemia can cause permanent brain injury, end-organ shock and cardiac arrest, and even mild or moderate hypoxemia contributes to increased mortality risk by decreasing resistance to infection and wound healing (Strachan & Noble, 2001).

Severe cases of COVID-19 are also characterized by hypoxemia and dyspnea (difficulty breathing) which can rapidly progress to respiratory failure (Berlin et al., 2020). These patients often require advanced life support measures including invasive mechanical ventilation, hospitalization in ICUs and even extra-corporeal membrane oxygenation (ECMO). During the COVID-19 pandemic, ventilators and ICU beds have become scarce resources with insufficient capacity in the hardest hit regions (Dar et al., 2020). As the COVID-19 pandemic continues to exact a heavy mortality toll with at least half a million deaths directly attributed to the disease in the United States alone and herd immunity by vaccination remains months away, it is important to find ways to manage these scarce resources and identify patients unable to maintain oxygen saturation without intervention. Clinically, an important decision point in the management of COVID-19 patients is determining whether the patient requires endotracheal intubation, a form of invasive ventilation (Berlin et al., 2020). Triage systems using monitoring of blood oxygenation to inform life support measures are tremendously useful for directing resource allocation and have been demonstrated to reduce mortality (Starr et al., 2020).

Given the host of negative clinical outcomes attributed to hypoxemia, identifying patients likely to experience acute hypoxemia in the near future would offer valuable opportunities for rapid intervention. Life support interventions ranging from supplemental oxygen to invasive ventilation prior to the onset of hypoxemia can mitigate or prevent the morbidity and mortality associated with hypoxemia (Strachan & Noble, 2001). Moreover, identifying patients not at imminent risk of hypoxemia represents an opportunity to conserve ventilators and ICU beds in the context of resource shortages arising from a global pandemic.

To this end, we present SWIFT (SpO_2_ <u>W</u>aveform <u>I</u>CU <u>F</u>orecasting <u>T</u>echnique), a neural network that predicts the blood oxygen saturation (SpO_2_) waveform for critically ill patients, 5 and 30 minutes in the future. SWIFT is unique for several reasons. First, SWIFT predicts both the occurrence and magnitude of hypoxemic events, and its prediction time horizon provides enough time for potential clinical interventions prior to acute desaturation events. Prior studies have made predictions on short time horizons (20 seconds to 5 minutes), leaving little room for potential clinical interventions (Elmoaqet et al., 2014; Erion et al., 2017; Ghazal et al., 2019; Lundberg et al., 2018). Moreover, most other attempts at hypoxemia prediction predict only a class value (hypoxemia vs. no hypoxemia, or mild hypoxemia vs. severe hypoxemia vs. no hypoxemia) rather than an actual SpO_2_ value (Elmoaqet et al., 2014; Erion et al., 2017; Ghazal et al., 2019). Clinically, there is a large difference between a transient dip in SpO_2_ to 91% versus an acute desaturation to 75% SpO_2_, though both would be considered hypoxemia. SWIFT recognizes this difference, hence providing important clinical information.

Second, SWIFT employs a Long Short-Term Memory (LSTM) architecture with only prior SpO_2_ values as inputs, hence allowing SWIFT to make predictions with limited, routinely acquired and readily available data. LSTM models are a type of recurrent neural network well-suited to modeling of time-series data that have shown promise in clinical applications (Che et al., 2018; Lin et al., 2019; Lipton et al., 2017). One prior study did use LSTM architectures with prior SpO_2_ values as inputs, but this model was limited to classification of timepoints as either hypoxemic or not with a 5 minute time horizon, and the total ROC-AUC was less than 0.75 (Erion et al., 2017). In contrast, other SpO_2_ prediction models have used complex, multifactorial data requiring extensive monitoring of patient vitals, demographic data, or ventilator settings (Ghazal et al., 2019; Lundberg et al., 2018). This limits their utility to only those patients for whom all of this data is readily available.

Third, SWIFT predicts more than 80% of all hypoxemic events (sensitivity) with positive predictive value (PPV) above 94% in test-sets of critically ill patients, and more than 60% of all hypoxemic events with PPV above 98% in a test-set of COVID-19 patients, across all timepoints for both the 5 minute and 30 minute time horizons. SWIFT also provides waveform predictions with an average mean squared error less than .0007 across all patient-stays. These results represent a marked improvement over recently published prediction algorithms. Auto-regressive models with PPV >90% have been limited to prediction time horizons less than 60 seconds (Elmoaqet et al., 2014), and ensemble-based machine learning models to classify hypoxemic events 5 minutes in the future were estimated to capture only 30% of hypoxemic events (Lundberg et al., 2018). To our knowledge, no other study has demonstrated waveform prediction.

Finally, SWIFT is highly generalizable. Though trained on only patients without COVID-19, it performs comparably on patients who received mechanical ventilation during their ICU stay and those who did not, and patients with and without a COVID-19 diagnosis. Other studies have been limited to specific groups such as pediatric patients on mechanical ventilation (Ghazal et al., 2019), orthopedic postoperative adult patients (Elmoaqet et al., 2014), or patients undergoing surgery in the operating room (Erion et al., 2017; Lundberg et al., 2018).

## Results

SWIFT is effective at predicting hypoxemia events (both occurrence and magnitude) across a variety of patient populations (Ventilated patients, non-ventilated patients, patients with COVID-19 diagnosis, critically ill patients without COVID-19 diagnosis). We show this through performance evaluation of SWIFT for prediction of hypoxemia at individual timepoints 5 and 30 minutes into the future, and evaluation of SWIFT for direct SpO_2_ waveform forecasting.

### SWIFT Overview

SWIFT utilizes an LSTM (Long Short-Term Memory) neural network architecture trained on the SpO_2_ waveforms from critically ill patients in the eICU database (Pollard et al., 2018) (Figure 1). The eICU database contains patients admitted to intensive care units across 208 United States hospitals in 2014 and 2015. We selected all unique ICU stays that include invasive ventilation at any point during the stay (1326 patient-stays), and then randomly sampled the same number of patient-stays without any use of invasive ventilation. We split the selected patient-stays into training and test sets: approximately 75%-25% split: 2000 training set patients (1000 each from the ventilator and non-ventilator groups) and 326 test set patients each in the ventilator group and non-ventilator group. Next, we excluded patient-stays with less than 5 hours of recorded vitals or entirely missing vitals. This left a total of 1837 training patient-stays (comprising both the ventilator and non-ventilator groups), and 310 and 288 patient-stays in the eICU ventilator and non-ventilator test sets, respectively (Figure 1a). A similar data processing procedure was used to obtain a test-set of patients from the Johns Hopkins JH-CROWN database, where all patients had COVID-19 (Figure 1b). We trained two different models, one which predicts hypoxemia 5 minutes in the future (SWIFT-5) and one which predicts hypoxemia 30 minutes in the future (SWIFT-30) (Figure 1c). Both models were trained using the SpO_2_ waveform sampled at 5 minute intervals with a causal moving average filter with a window of 5 time points (4 previous time points, plus current time point). For SWIFT-30, the data was down-sampled to an interval of 30 minutes before training. Both models take the two previous timepoints of data (10 minutes prior data for SWIFT-5, 60 minutes prior data for SWIFT-30) as inputs and predict SpO_2_ value for the next time point. We used these predictions to forecast the exact SpO_2_ waveform, and to classify individual timepoints as hypoxic events based on a threshold of 92% SpO_2_.

**Figure 1:**
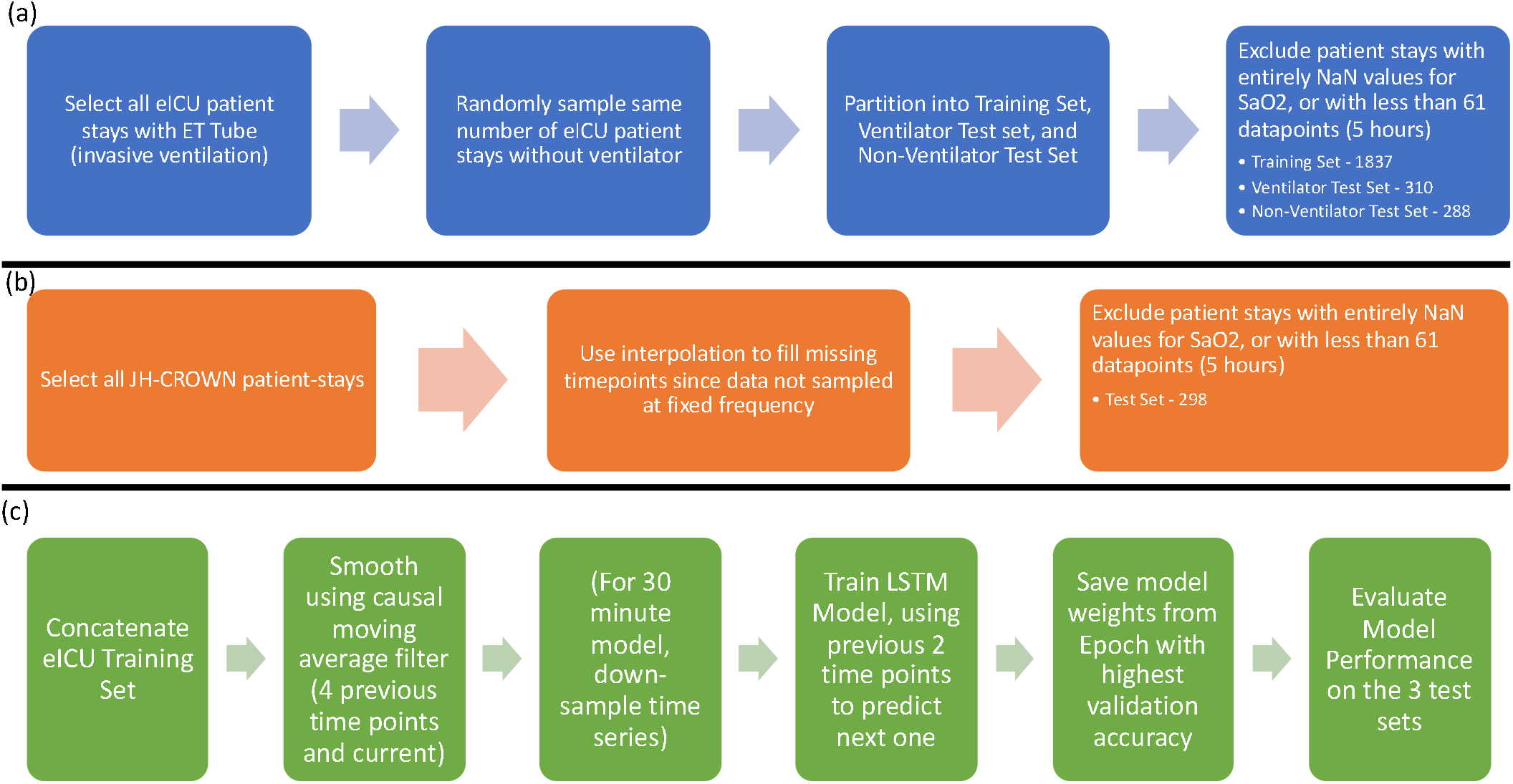
Schematic illustrating data processing for (a) eICU patients and (b) JH-CROWN patients, and (c) model training and testing pipelines

We considered 92% SpO_2_ to be the threshold for hypoxemia, as SpO_2_ below 92% has been shown to be associated with adverse events in a broad population of outpatient adults with pneumonia (Majumdar et al., 2011), and is the low end of the National Institutes of Health’s recommended SpO_2_ target range for COVID-19 patients (Shenoy et al., 2020). Moreover, 92% SpO_2_ is between the World Health Organization’s hypoxemia treatment threshold (94%) and clinical emergency threshold (90%) (World Health Organization, 2011), and has been used as the hypoxemia cut-off in other prediction studies (Ghazal et al., 2019; Lundberg et al., 2018).

We demonstrate the SpO_2_ waveform and hypoxemia prediction capabilities of SWIFT on three different test sets of patient-stays: patient-stays with and without the use of invasive ventilation at any point in the stay from the eICU database, and a test set of critically ill COVID-19 patients from the JH-CROWN database. While the eICU database consists of critically ill patients with a variety of diagnoses, the JH-CROWN database consists of patients specifically diagnosed with and admitted to hospital for COVID-19 at a single academic center and its affiliated hospitals in 2020.

We hypothesized that patients requiring invasive ventilation at some point in their ICU stay would have different profiles with respect to hypoxemia than those never requiring invasive ventilation. A chi-squared test of independence between use of mechanical ventilation and number of hypoxemic time-points in the dataset was statistically significant for all groups (eICU 30 minute time-series, p=1.4e-303; eICU 5 minute time-series, p=0.0), hence motivating our treatment of eICU patient-stays utilizing invasive ventilation at any point as a distinct test set from those patient-stays never utilizing invasive ventilation. We did not split the JH-CROWN test-set into ventilator and non-ventilator test-sets since all patients had the same diagnosis and the overwhelming majority were on ventilators, in contrast to the patients in the eICU database. Importantly, the different patient populations studied vary dramatically with respect to frequency of hypoxic events, duration of available time-series data, and reason for hospital admission (Table 1), yet our models are effective at recapitulating the patient’s SpO_2_ waveform over time and predicting hypoxemic events across these diverse patient populations.

**Table 1:** Summary of patient characteristics used in model testing. (Note: Demographic information on age, gender, and race only available for 294 of 298 patients in the JH-CROWN dataset)

| Characteristics | eICU Ventilator – 30 minutes | eICU non-Ventilator – 30 minutes | JH CROWN– 30 minutes | eICU Ventilator – 5 minutes | eICU non-Ventilator – 5 minutes | JH CROWN– 5 minutes |
| --- | --- | --- | --- | --- | --- | --- |
| # Patient Stays | 310 | 288 | 298 | 310 | 288 | 298 |
| Median Hypoxic events/patient stay | 9 | 1 | 79.5 | 55 | 4 | 481 |
| Number patient-stays with No Hypoxic events | 64 | 138 | 10 | 53 | 123 | 10 |
| Number patient-stays with all Hypoxic events | 4 | 1 | 1 | 3 | 0 | 1 |
| Median number of timepoints per patient-stay | 133 | 79.5 | 1083 | 798 | 477 | 6497 |
| Median Age at patient-stay | 62.0 (on admission) | 65.0 (on admission) | 60.98 (on Dec 15, 2020 assuming 365 days/year) | 62.0 (on admission) | 65.0 (on admission) | 60.98 (on Dec 15, 2020 assuming 365 days/year) |
| % patient-stays with Female patients | 42.9% | 49.3% | 41.2% | 42.9% | 49.3% | 41.2% |
| % patient-stays with White/Caucasian patients | 81.3% | 76.7% | 26.2% | 81.3% | 76.7% | 26.2% |
| Admissions Diagnosis | Top 3:<br>(1) CABG alone, coronary artery bypass grafting (42 stays)<br>(2) Cardiac arrest (24 stays)<br>(3) Emphysema/bronchitis (15 stays) | Top 3:<br>(1) Not recorded (15 stays)<br>(2) Sepsis, pulmonary (14 stays)<br>(3) Rhythm disturbance (13 stays) | All patients with COVID-19 | Top 3:<br>(1) CABG alone, coronary artery bypass grafting (42 stays)<br>(2) Cardiac arrest (24 stays)<br>(3) Emphysema/bronchitis (15 stays) | Top 3:<br>(1) Not recorded (15 stays)<br>(2) Sepsis, pulmonary (14 stays)<br>(3) Rhythm disturbance (13 stays) | All patients with COVID-19 |

### SWIFT effectively predicts hypoxic events 5 and 30 minutes in the future

We used SWIFT on the SpO_2_ time-series of each patient-stay in the three test sets to predict, based on the prior two time points, whether each time-point was expected to be a hypoxemic event or not. Averaging across the value for each patient-stay, SWIFT-5 and SWIFT-30 both achieved mean accuracy greater than 96% for both the eICU and JH-CROWN patient-stays (Figure 2a), mean sensitivity greater than 73% for eICU patient-stays and greater than 59% for JH-CROWN patient-stays (Figure 2b), mean specificity greater than 99% for both the eICU and JH-CROWN patient-stays (Figure 2c) and mean PPV greater than 84% for eICU-patient stays and greater than 97% for JH-CROWN patient-stays (Figure 2d). The quality of predictions made for COVID patients is comparable to that of the predictions made on patients without COVID, which is notable given that SWIFT was trained exclusively on non-COVID patients. Interestingly, SWIFT-30 demonstrates performance comparable to SWIFT-5, despite the much larger prediction time horizon. While there are a few patient-stays for whom sensitivity and PPV are low (<50%), by and large these models predict hypoxemic events effectively. In particular, the high PPV achieved illustrates the utility of these models for identifying true hypoxemic events.

**Figure 2:**
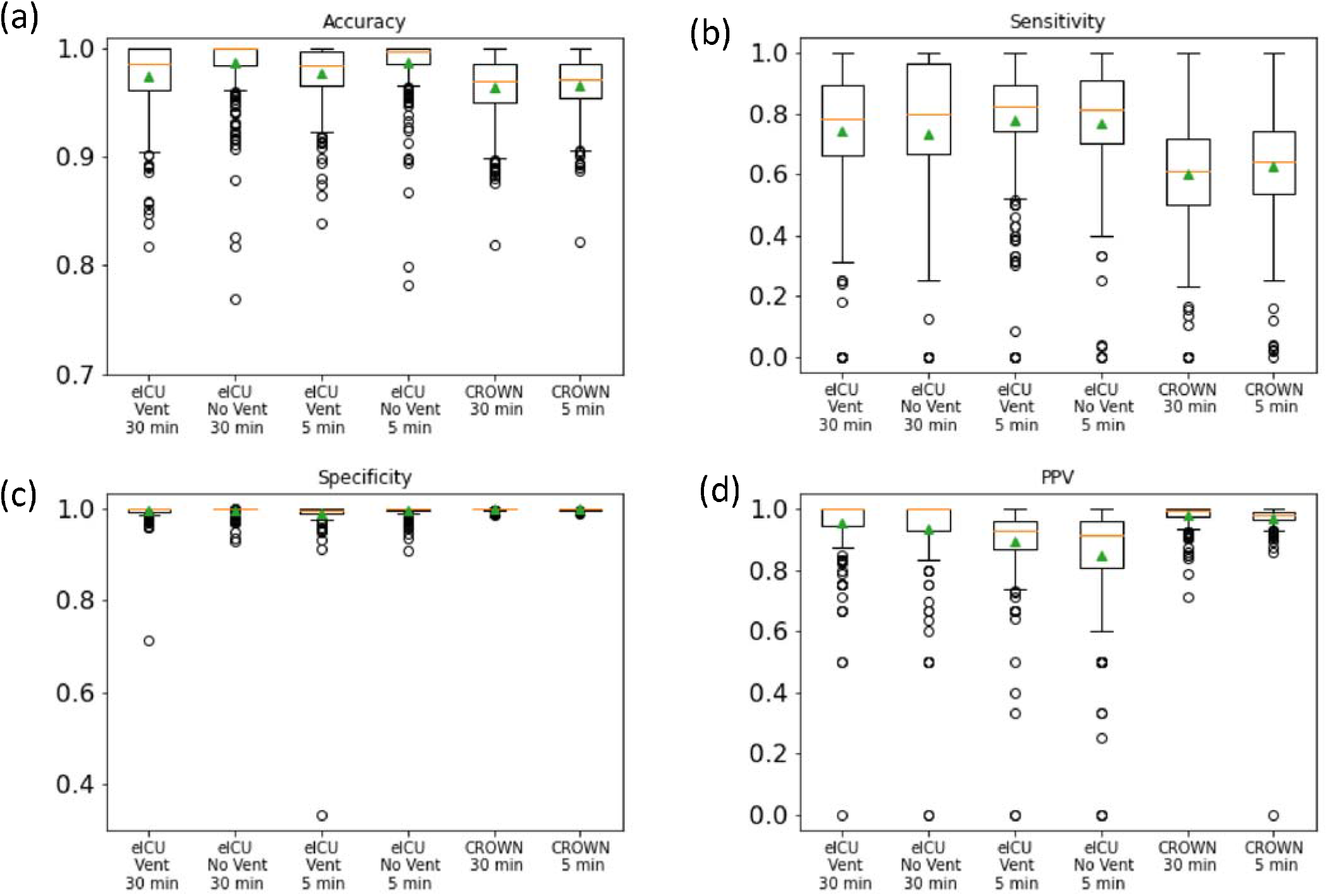
(a) Accuracy, (b) Sensitivity, (c) Specificity, and (d) PPV for SWIFT-5 and SWIFT-30 tested on eICU patients with and without ventilators, and JH-CROWN patients with COVID-19. In each box-and-whisker plot, the individual datapoints come from evaluation of the model on each of the test-set patients. The box extends from Q1 to Q3. The orange line represents the median value and the green triangle represents the mean value. The upper whisker extends to the highest value below Q3+1.5*(Q3-Q1), and the lower whisker extends to the lowest value below Q1-1.5*(Q3-Q1). Points beyond the whiskers are considered outliers.

Moreover, when all timepoints are aggregated across patient-stays, SWIFT classifies hypoxic events with remarkable success (Figure 3). Across all time-points in all test-sets for both SWIFT-5 and SWIFT-30 the False Positive rate is less than 0.65% and the False Negative Rate is less than 4%. While the datasets are imbalanced with far less hypoxic timepoints than non-hypoxic events, the high PPV and sensitivity demonstrate that SWIFT accurately classifies hypoxic timepoints. Both SWIFT-5 and SWIFT-30 represent a substantial improvement over the current state-of-the art method, Prescience, which uses machine learning to predict hypoxic events in a 5 minute window, with a binary classifier (Lundberg et al., 2018). Prescience predicts 44% of hypoxic events (recall or sensitivity) with a precision (or PPV) of 30%. For a PPV of 70%, Prescience’s sensitivity falls to 9%. In contrast, SWIFT uses a waveform forecasting approach to achieve PPV above 94% and sensitivity above 80% across all timepoints in the eICU test sets, and PPV above 98% and sensitivity above 60% across all timepoints in the JH-CROWN test set.

**Figure 3:**
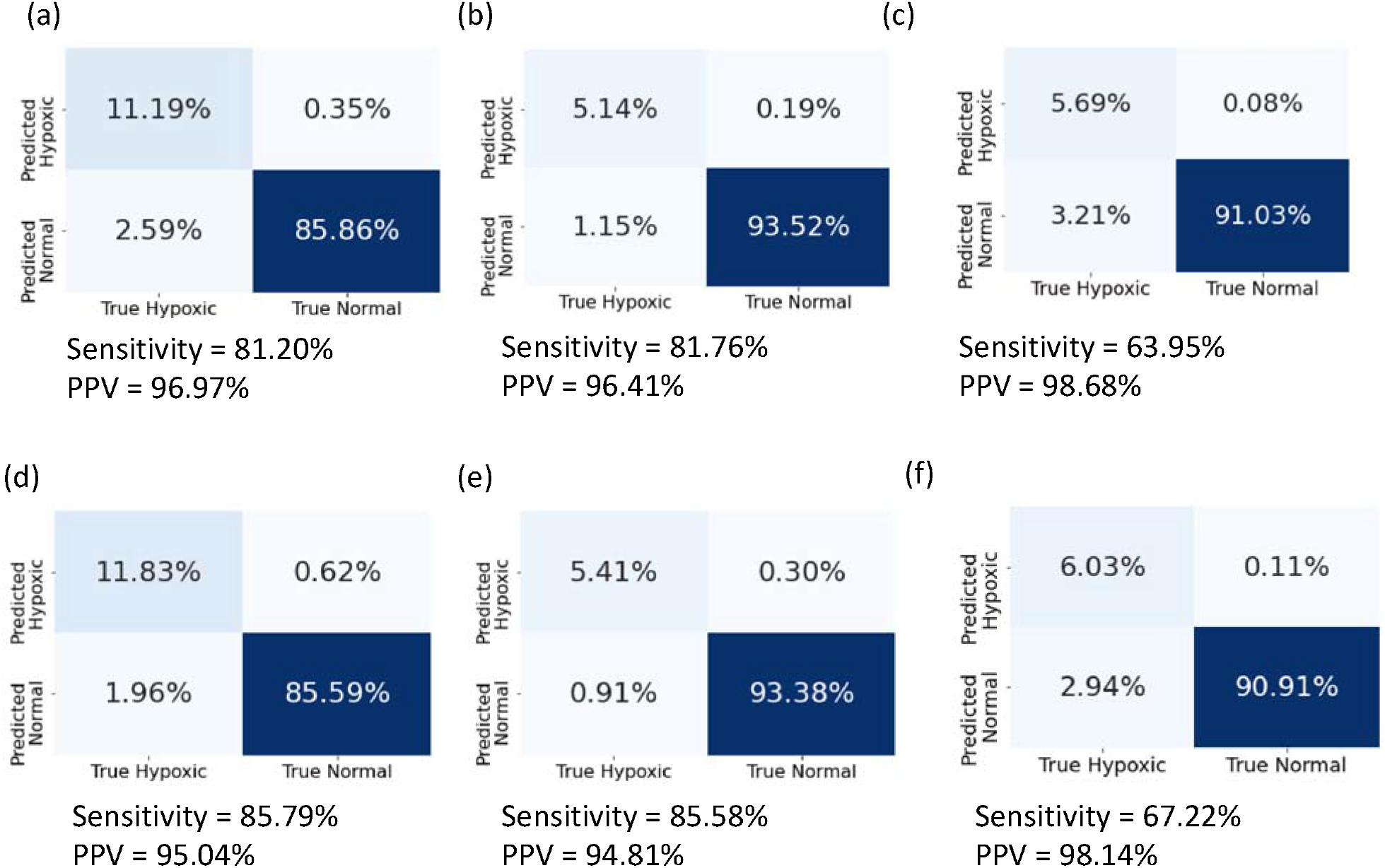
Confusion matrices aggregated across all timepoints for all patients for (a) eICU Ventilator – 30 minute, (b) eICU No ventilator – 30 minute, (c) JH-CROWN – 30 minute, (d) eICU Ventilator – 5 minute, (e) eICU No ventilator – 5 minute, (f) JH-CROWN– 5 minute

### SWIFT effectively predicts the SpO_2_ waveform for individual patients

We used SWIFT to predict the SpO_2_ time-series of each patient-stay in the three test sets in order to determine the magnitude of potential hypoxic events and the overall time-evolution of transient hypoxic events. On average across all patient-stays, SWIFT-5 and SWIFT-30 both achieved mean-squared-error (MSE) from the true patient time-series of less than .0007 (Figure 4a), and a Pearson correlation coefficient with the true patient time-series of greater than .95 (Figure 4b). This indicates that the waveform predictions used to predict hypoxic events are extremely close to the true waveforms. Figure 5 shows examples of SWIFT-30’s best and worst by waveform predictions by MSE (Figure S1 contains the same information for SWIFT-5), qualitatively demonstrating the accuracy with which SWIFT recapitulates SpO_2_ waveform.

**Figure 4:**
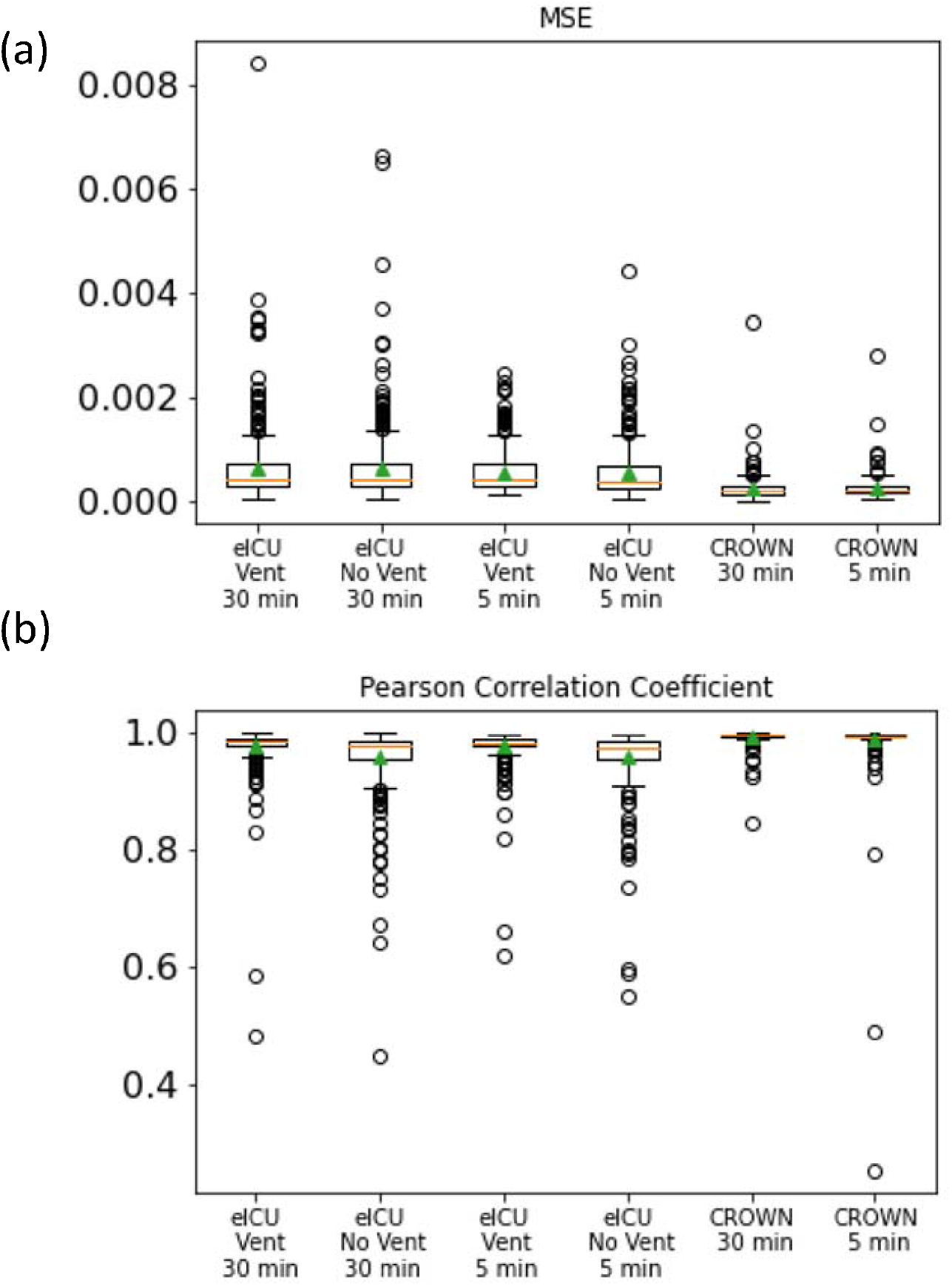
(a) Mean-Squared Error and (b) Pearson Correlation Coefficient for waveform predictions for eICU and JH-CROWN patients. In each box-and-whisker plot, the box extends from Q1 to Q3. The orange line represents the median value and the green triangle represents the mean value. The upper whisker extends to the highest value below Q3+1.5*(Q3-Q1), and the lower whisker extends to the lowest value below Q1-1.5*(Q3-Q1). Points beyond the whiskers are considered outliers.

**Figure 5:**
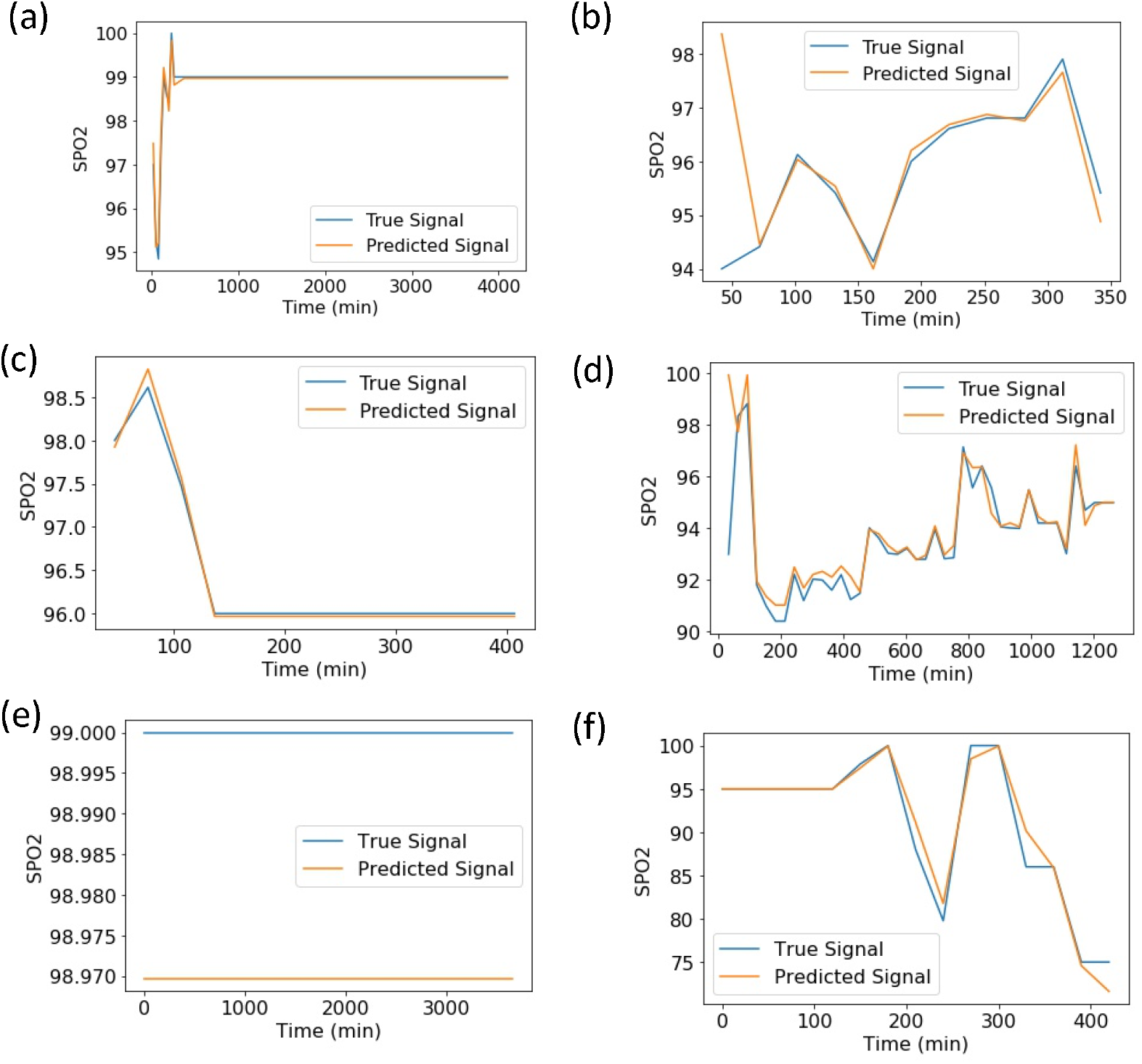
Examples of the best and worst fits by MSE for SWIFT-30 model tested on (a) eICU Ventilated patients – best fit, (b) eICU Ventilated patients – worst fit, (c) eICU Non-Ventilated patients – best fit, (d) eICU Non-Ventilated patients – worst fit, (e) JH-CROWN patients – best fit, (f) JH-CROWN patients – worst fit

This waveform prediction allows for the forecasting of the magnitude of hypoxic events rather than their occurrence alone. This may have implications for patient management, especially in the context of limited ICU beds and shortages of ventilation machinery during the COVID-19 pandemic (Dar et al., 2020).

## Discussion

SWIFT is a Long Short-Term Memory neural network model capable of predicting the magnitude and occurrence of hypoxemic events 5 and 30 minutes in the future, using only prior SpO_2_ values. We tested SWIFT on three different test sets of ICU patient-stays, including patients both requiring and not requiring mechanical ventilation during their ICU stay, and patients with and without COVID-19. Across all time points in these test-sets, SWIFT predicts more than 80% of all hypoxemic events (sensitivity) with PPV above 94% in test-sets of critically ill patients, and more than 60% of all hypoxemic events with PPV above 98% in test-sets of COVID-19 patients, for both the 5 minute and 30 minute time horizons. Additionally, SWIFT-5 and SWIFT-30 accurately predicted SpO_2_ waveforms for each patient-stay with an average MSE below .0007 and an average Pearson’s correlation coefficient greater than .95.

SWIFT may be especially useful in the context of the COVID-19 pandemic or future similar pandemics with high numbers of patients experiencing hypoxemia and limited supplies of ventilators and ICU beds. Strategies to reduce the demand for mechanical ventilation have been identified as a priority for resource management during the pandemic (Dar et al., 2020). To this end, SWIFT can help identify patients likely to experience imminent hypoxemic events versus patients likely to remain stable and offer insights into the magnitude of the potential hypoxemic event. This can enable the increased management of patients off of ventilators, and if needed, offer another data point to be used in the triaging of patients for therapy.

Beyond the COVID-19 pandemic, SWIFT could be easily deployed in real time, in low-resource settings without access to complex clinical informatics or large amounts of memory storage. Since SWIFT’s only model inputs are two previous values of SpO_2_, the barriers to use are minimal. SpO_2_ can be assessed using simple, non-invasive pulse oximeters. Pulse oximetry is nearly ubiquitous in hospitals and critical care units in the developed world, and substantial effort has been dedicated to increasing the use of pulse oximetry in low resource settings (Herbert & Wilson, 2012). Given the existing need for hypoxemia monitoring in low and middle income countries and challenges in access to oxygen therapy, SWIFT’s predictive capabilities can play a crucial role in identifying patients likely to experience hypoxemic events and in informing resource allocation decisions (Lam et al., 2020).

Moreover, SWIFT provides benefits by waveform prediction of SpO_2_ rather than only binary classification of events as provided by other existing models (Erion et al., 2017; Ghazal et al., 2019; Lundberg et al., 2018). Studies have demonstrated that pulse oximetry has high levels of false alarms, often for clinically insignificant reasons such as patient movement or skin condition, which can contribute to alarm fatigue (Nguyen et al., 2018; Winters et al., 2018). Alarm fatigue may lead to slower or absent responses to truly dangerous events (Sendelbach & Funk, 2013). Since SWIFT provides a prediction of SpO_2_ magnitude as much as 30 minutes in the future, minor anticipated hypoxemic events can be distinguished from more severe ones, with sufficient time horizon to allow for decision making on this basis. While SWIFT cannot correct for errors in SpO_2_ readings caused by the pulse oximetry device, it can anticipate transient dips hence preventing unnecessary response to transient SpO_2_ dips that may occur for clinically insignificant reasons. This may have a beneficial effect on controlling the phenomena of alarm fatigue.

Importantly, SWIFT generalizes well across patient groups. We did not observe substantial differences in model performance between SWIFT-5 and SWIFT-30, nor between predictions made on ventilated vs. non-ventilated patients and COVID-19 vs. generally critically ill patients. The one exception was sensitivity, when aggregated across all timepoints – in the eICU test sets, more than 80% of hypoxemic events were detected as compared to more than 60% in the JH-CROWN test sets. This is unsurprising given that SWIFT was trained exclusively on non-COVID-19 patients, and the JH-CROWN database consists only of COVID-19 patients. Notably, the lung damage from SARS CoV-2 infection appears more severe than that from Acute Respiratory Distress Syndrome (ARDS) secondary to most other etiologies, and we are still in the early stages of understanding COVID-19 disease mechanistically. This difference in degree of lung damage may contribute to the performance differences.

Moreover, these predictions appear to be generalizable across hospitals and dates (the eICU database comprises patient-stays from 208 ICUs in 2014 and 2015, whereas the JH-CROWN database consists of patients from one medical center in 2020). Our test-sets contained male and female patients in roughly equal proportions, a range of admissions diagnoses, and substantial numbers of patients with non-white ethnicities (Table 1).

However, one limitation of SWIFT is that it was trained and tested primarily on older, critically ill patients. The median age of patients in each test-set was between 60 and 65 years old, and all data came from critically ill patients. Hypoxemia is a consideration in much younger patients as well, and future work will be needed to evaluate SWIFT-5 and SWIFT-30 on younger patients, or to train new models with additional data. A second limitation is that we did not train race-specific models. Recent work has shown that occult hypoxemia (low arterial oxygen saturation despite a pulse oximetry measurement between 92% and 96%) occurs far more frequently in Black patients than White patients (Sjoding et al., 2020). For this reason, there is racial bias in interpretation of SpO_2_ values, which may not be well captured by our models (though our test sets are racially diverse; the JH-CROWN test sets have ∼75% non-White patients). Regardless, SWIFT currently demonstrates high potential utility for simple, real-time prediction of hypoxemic events (occurrence and magnitude) 5 and 30 minutes in the future without the use of complex clinical informatics. As part of a clinical decision support system, SWIFT has the potential to inform the management of critically ill patients at risk for hypoxemia, including COVID-19 patients.

## Methods

### Data Selection

First, we selected all patient ICU stays with mechanical ventilation at some point during the ICU stay from the eICU database (n=1326) (Pollard et al., 2018). The eICU database consists of critically ill patients treated in 208 intensive care units across the United States in 2014 and 2015. We defined ICU stays with mechanical ventilation as distinct *patientUnitStayID* identifiers for which a respiratory chart entry included phrases similar to ET TUBE, ETT, Endotracheal, Trach, or Tracheostomy. Then, we randomly selected 1326 *patientUnitStayID* identifiers from those without indication of mechanical ventilation. We partitioned the first 1000 *patientUnitStayID* identifiers from the mechanical ventilation and no mechanical ventilation groups into a training set (n=2000), and the last 326 from each group into two eICU test sets (eICU mechanical ventilation n=326, eICU no mechanical ventilation n=326). Then, we queried the vital signs time-series for each of these ICU stays, and excluded any ICU stays without corresponding SpO_2_ data recorded. This left 1933 stays in the training set, 326 stays in the mechanical ventilation test set, and 311 stays in the no mechanical ventilation test set. Since it is possible for a patient in the eICU database to have multiple ICU stays, we took the additional step of removing all ICU stays from the test sets for which that patient had a different ICU stay in the training set. This ensured that there was no overlap in patients between the train and test sets despite being unique ICU stays. This left 1933 stays (corresponding to 1859 patients) in the training set, 317 stays (corresponding to 285 patients) in the mechanical ventilation test set, and 311 stays (corresponding to 306 patients) in the no mechanical ventilation test set.

Second, all patient stays from the JH-CROWN database up to December 15, 2020 were selected (n=301). The JH-CROWN database consists of COVID-19 patients seen in any Johns Hopkins Medical Institution facility with confirmed or suspected COVID-19. Each patient-stay in the JH-CROWN database corresponds to a unique patient (n=301). Data extraction was performed using PostgreSQL, and the Python libraries psycopg2 and pandas (McKinney, 2010).

Finally, those patients with entirely blank values for SpO_2_ were excluded. The eICU database contains vital signs recorded at 5 minute intervals, whereas the JH-CROWN database records vital signs at variable frequency. Therefore, the data in the JH-CROWN database was interpolated to 5 minute intervals by replacing blank values of SpO_2_ with the last valid observation. In the patients selected from the JH-CROWN database, the median time between observations was 25 minutes. If the first SpO_2_ value was missing, it was backfilled with the first available SpO_2_ value. Finally, those time series with less than 61 datapoints (5 hours) were excluded. This left 1837 patient-stays for model training, and 310, 288, and 298 patient-stays in the eICU Mechanical Ventilation, eICU No Mechanical Ventilation and JH-CROWN test sets respectively (Fig S2, S3).

### Data Preparation

All SpO_2_ values were transformed using the following equation:

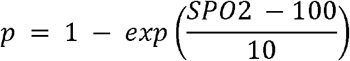

This transformed value was chosen to magnify differences between SpO_2_ values close to 100%. Next, a causal moving average filter with a window of 5 was applied to each patient’s transformed SpO_2_ waveform (ie, the SpO_2_ values at the previous 4 timepoints and the current timepoint were averaged together). We chose this data smoothing technique since it is causal, meaning that it can be applied in real time, and it reduces transient noise hence providing a less noisy signal more suitable for clinical decision making. Other studies of hypoxemia prediction have also applied averaging filters to time-series data prior to prediction (Elmoaqet et al., 2014; Lundberg et al., 2018). The time series for each patient was then down-sampled to 30 minute frequency for use with SWIFT-30 model which predicts SpO_2_ 30 minutes in the future. For SWIFT-5, which predicts SpO_2_ 5 minutes in the future, no changes were made.

Finally, the smoothed time series data for each patient was rearranged into an input vector X, and output vector Y where *p*_*n*_ is SpO_2_ at timestep n:

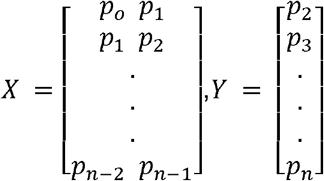

Finally, in the training set, all X vectors were concatenated and all Y vectors were concatenated to create one training set input vector and one training set output vector to be used in model training. In the 4 test sets, the patient-level input and output vectors were maintained to be used for model testing. Data preparation was performed in Python using standard data science libraries (Harris et al., 2020; McKinney, 2010; Pedregosa et al., 2011).

### Model Training

A 3-fold cross validation procedure was used for hyperparameter optimization on the training data to evaluate 2 different LSTM model architectures (a deep architecture with 5 LSTM hidden layers and a shallow architecture with 2 LSTM hidden layers; both models had a Batch Normalization input layer and a Dense 1 neuron output layer and contained Dropout layers to prevent overfitting) and 3 different learning rates (ADAM optimizer with learning rates .001, .01 and .1). For both SWIFT-5 and SWIFT-30, the shallow architecture with learning rate .001 demonstrated the lowest average MSE across folds. This architecture was then used to re-train the final models on the full training set. The models were trained for 100 epochs with a random 10% validation set at each epoch. To prevent overfitting, the model weights at the epoch with lowest validation loss were used for the final SWIFT-5 and SWIFT-30 models. All model training was performed using the TensorFlow and Keras libraries in Python (Abadi et al., n.d.; Chollet, 2015).

### Model Testing

SWIFT-5 and SWIFT-30 were used to predict the transformed SpO_2_ waveform for each individual patient-stay in each of the three test sets. Then, the mean-squared-error and Pearson’s correlation coefficient were calculated between the true and predicted waveform for each patient-stay. Pearson’s correlation coefficient could not be calculated for 1 patient-stay in the JH-CROWN test set since the time series was constant and the correlation coefficient was undefined. Next, each time point was classified as hypoxemic or not based on a threshold of SpO_2_ 92% (transformed SpO_2_ .55067). Each prediction was also checked against the same threshold, and the sensitivity, specificity, accuracy, and PPV were calculated for each patient-stay time series. Sensitivity was not calculated for those patient-stays with no hypoxic events; Specificity was not calculated for those patient-stays with all hypoxic events; PPV was not calculated for those patient-stays for which no predictions were positive for hypoxemia, since these values are undefined in these cases. Finally, all timepoints in each test set were aggregated, and the false positive, true positive, false negative, and true negative rates were calculated for each test set.

## Data Availability

Some of the data utilized for this publication were part of the JH-CROWN: The COVID PMAP Registry which is based on the contribution of many patients and clinicians. The JH-CROWN Database is described here: https://ictr.johnshopkins.edu/coronavirus/jh-crown/
The JH-CROWN data is not publicly available due to IRB restrictions.
This study was approved by The Johns Hopkins School of Medicine IRB 00251922: RAPID Ventilator Modeling for Management of Covid 19 (SARS CoV-2) induced ARDS.
The other data set used is the eICU Collaborative Research Database. It is available here with access requirements: https://eicu-crd.mit.edu/gettingstarted/access/

https://ictr.johnshopkins.edu/coronavirus/jh-crown/

https://eicu-crd.mit.edu/gettingstarted/access/

## Acknowledgements

Stephen Granite assisted with accessing the data sets.

## Data/Code Availability

Some of the data utilized for this publication were part of the JH-CROWN: The COVID PMAP Registry which is based on the contribution of many patients and clinicians. The JH-CROWN Database is described here: https://ictr.johnshopkins.edu/coronavirus/jh-crown/

The JH-CROWN data is not publicly available due to IRB restrictions.

This study was approved by The Johns Hopkins School of Medicine IRB 00251922: RAPID Ventilator Modeling for Management of Covid 19 (SARS CoV-2) induced ARDS.

The other data set used is the eICU Collaborative Research Database. It is available here with access requirements: https://eicu-crd.mit.edu/gettingstarted/access/

There is no need for IRB approval to use eICU, as it is a publicly available de-identified database and therefore meets the conditions for exemption as human subjects research.

All code was written in Python including the use of scipy, scikitlearn, numpy, keras and tensorflow libraries. The code used to train and evaluate the models is available here: https://github.com/JHU-Winslow-Lab/hypoxemia-pred

## Author Contributions

JLG, RLW and SVS initiated the study. BDW, SVS and RLW wrote and submitted the IRB and data use agreements. AVA and SNB performed data cleaning and processing. AVA constructed the models and performed the analysis. BDW provided clinical consultation. JLG, SNB, RLW, and SVS provided consultation on the methods and analysis. AVA wrote the paper and JLG, SNB, BDW, SVS and RLW provided manuscript feedback.

---

**Figure S1:**
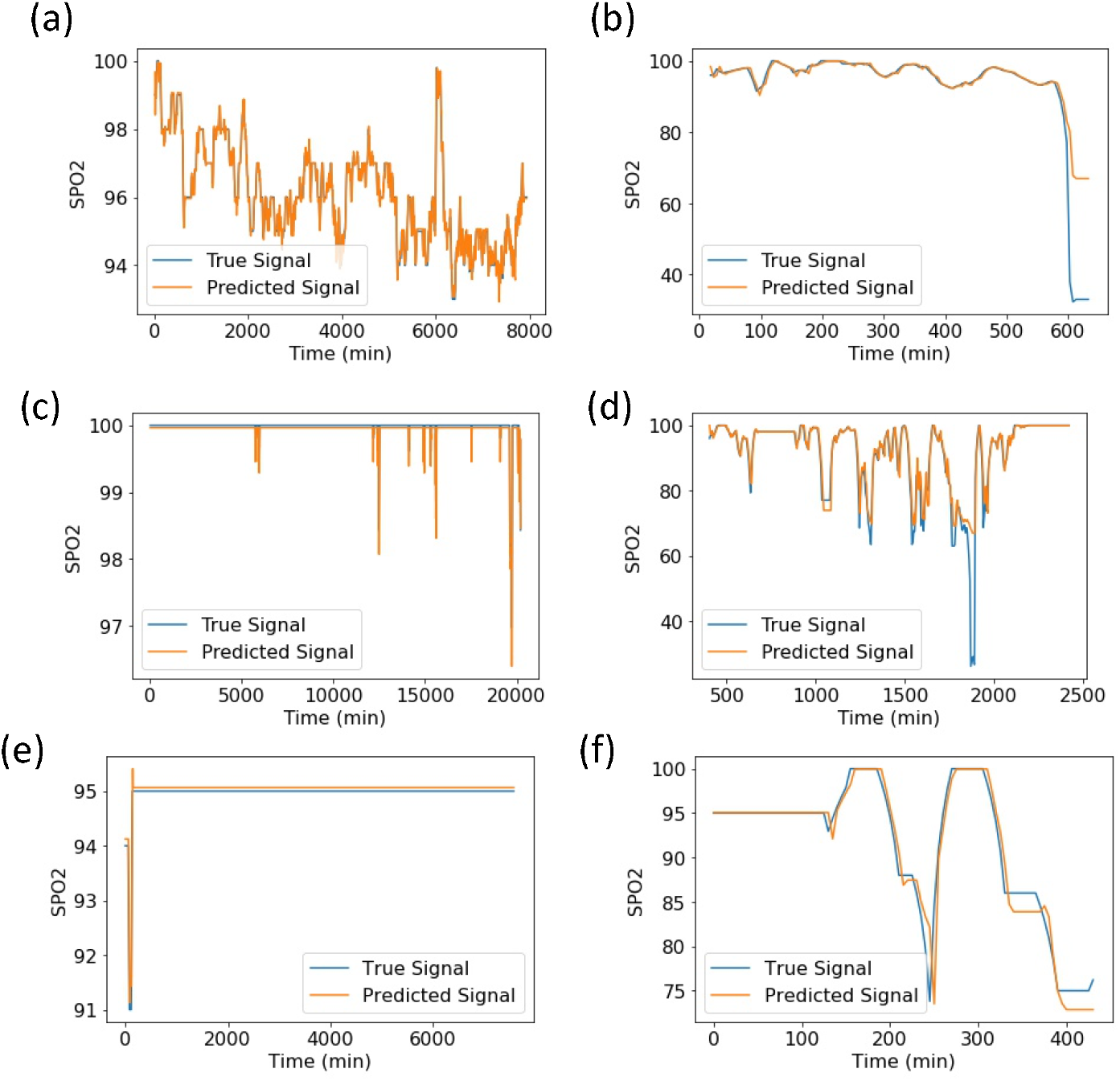
Examples of the best and worst fits by MSE for SWIFT-5 model tested on (a) eICU Ventilated patients – best fit, (b) eICU Ventilated patients – worst fit, (c) eICU Non-Ventilated patients – best fit, (d) eICU Non-Ventilated patients – worst fit, (e) JH-CROWN patients – best fit, (f) JH-CROWN patients – worst fit

**Fig S2:**
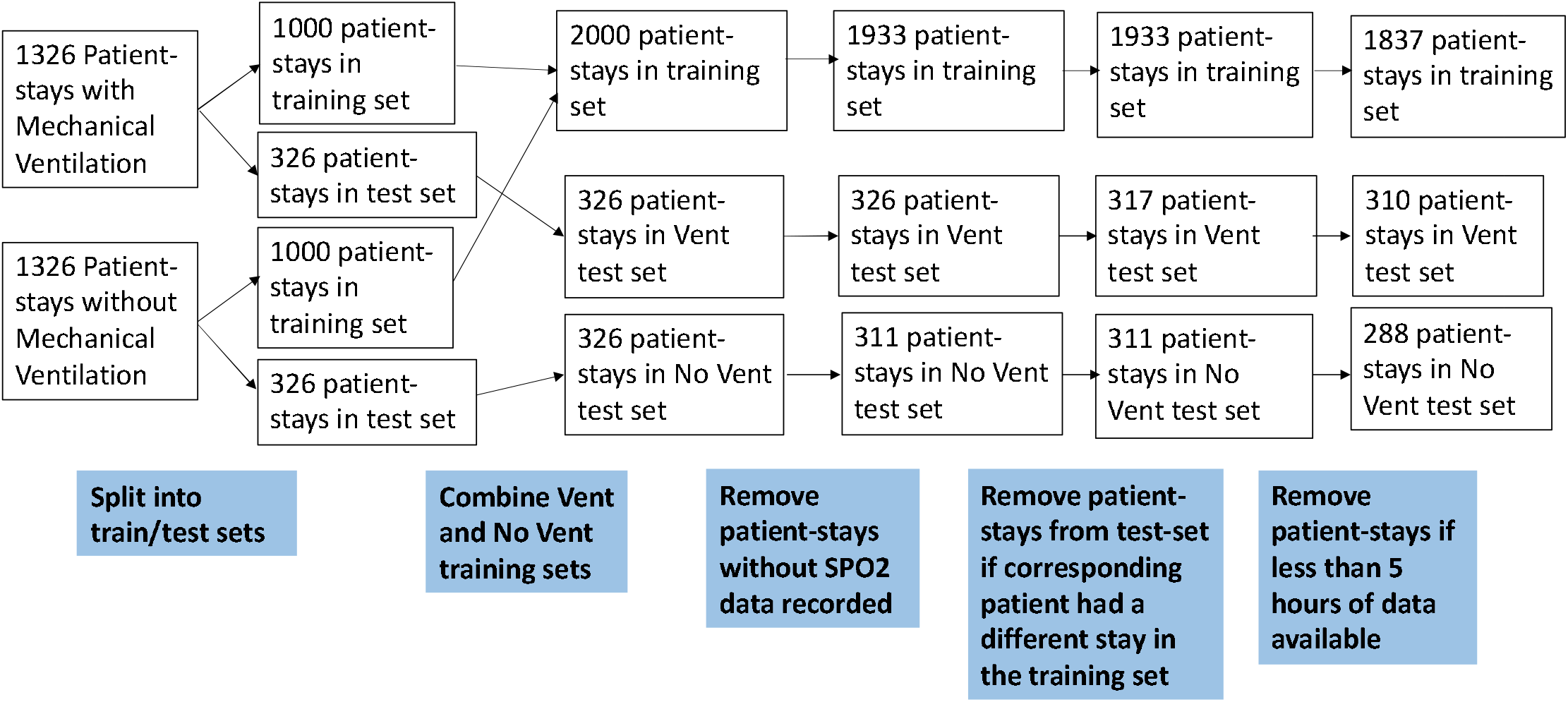
Inclusion/Exclusion Diagram for patient-stays from eICU Database

**Fig S3:**
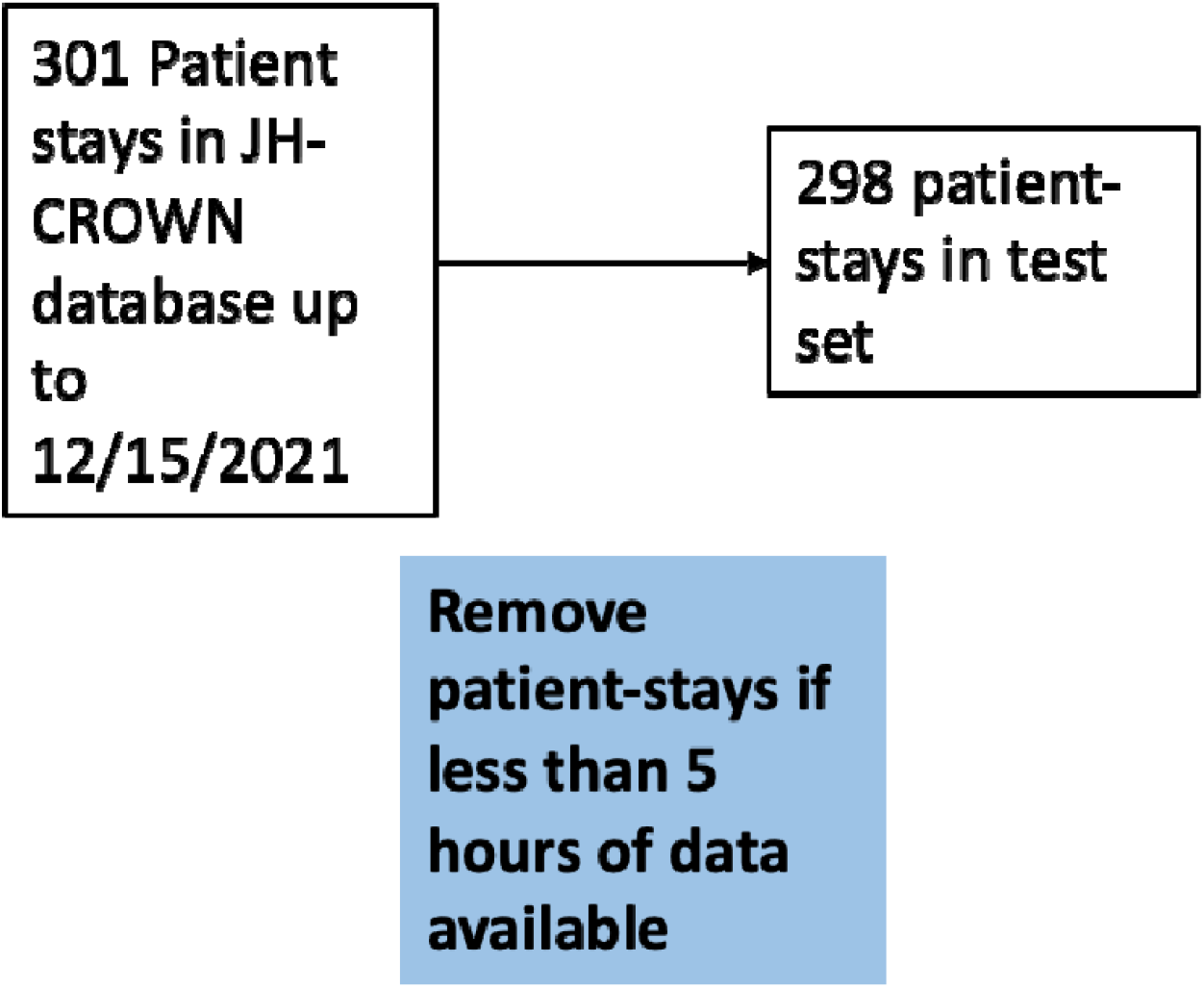
Inclusion/Exclusion Diagram for patient-stays from JH-CROWN Database

## References

Abadi, M., Agarwal, A., Barham, P., Brevdo, E., Chen, Z., Citro, C., Corrado, G. S., Davis, A., Dean, J., Devin, M., Ghemawat, S., Goodfellow, I., Harp, A., Irving, G., Isard, M., Jia, Y., Jozefowicz, R., Kaiser, L., Kudlur, M., … Zheng, X. (n.d.). TensorFlow: Large-Scale Machine Learning on Heterogeneous Distributed Systems. 19.

Berlin, D. A., Gulick, R. M., & Martinez, F. J. (2020). Severe Covid-19. The New England Journal of Medicine. https://doi.org/10.1056/NEJMcp2009575

Che, Z., Purushotham, S., Cho, K., Sontag, D., & Liu, Y. (2018). Recurrent Neural Networks for Multivariate Time Series with Missing Values. Scientific Reports, 8(1), 6085. https://doi.org/10.1038/s41598-018-24271-9

Chollet, F. (2015). Keras. \url{https://keras.io}

Dar, M., Swamy, L., Gavin, D., & Theodore, A. (2020). Mechanical Ventilation Supply and Options for the COVID-19 Pandemic: Leveraging All Available Resources for a Limited Resource in a Crisis. Annals of the American Thoracic Society. https://doi.org/10.1513/AnnalsATS.202004-317CME

Elmoaqet, H. Tilbury, D. M., & Ramachandran, S. K. (2014). Evaluating predictions of critical oxygen desaturation events. Physiological Measurement, 35(4), 639–655. https://doi.org/10.1088/0967-3334/35/4/639

Erion, G., Chen, H., Lundberg, S. M., & Lee, S.-I. (2017). Anesthesiologist-level forecasting of hypoxemia with only SpO2 data using deep learning. ArXiv:1712.00563 [Cs, Stat]. http://arxiv.org/abs/1712.00563

Ghazal, S., Sauthier, M., Brossier, D., Bouachir, W., Jouvet, P. A., & Noumeir, R. (2019). Using machine learning models to predict oxygen saturation following ventilator support adjustment in critically ill children: A single center pilot study. PloS One, 14(2), e0198921. https://doi.org/10.1371/journal.pone.0198921

Harris, C. R., Millman, K. J., van der Walt, S. J., Gommers, R., Virtanen, P., Cournapeau, D., Wieser, E., Taylor, J., Berg, S., Smith, N. J., Kern, R., Picus, M., Hoyer, S., van Kerkwijk, M. H., Brett, M., Haldane, A., del Río, J. F., Wiebe, M., Peterson, P., … Oliphant, T. E. (2020). Array programming with NumPy. Nature, 585(7825), 357–362. https://doi.org/10.1038/s41586-020-2649-2

Herbert, L. J., & Wilson, I. H. (2012). Pulse oximetry in low-resource settings. Breathe, 9(2), 90–98. https://doi.org/10.1183/20734735.038612

Lam, F., Subhi, R., Houdek, J., Schroder, K., Battu, A., & Graham, H. (2020). The prevalence of hypoxemia among pediatric and adult patients presenting to healthcare facilities in low-and middle-income countries: Protocol for a systematic review and meta-analysis. Systematic Reviews, 9(1), 67. https://doi.org/10.1186/s13643-020-01326-5

Lin, Y.-W., Zhou, Y., Faghri, F., Shaw, M. J., & Campbell, R. H. (2019). Analysis and prediction of unplanned intensive care unit readmission using recurrent neural networks with long short-term memory. PLOS ONE, 14(7), e0218942. https://doi.org/10.1371/journal.pone.0218942

Lipton, Z. C., Kale, D. C., Elkan, C., & Wetzel, R. (2017). Learning to Diagnose with LSTM Recurrent Neural Networks. ArXiv:1511.03677 [Cs]. http://arxiv.org/abs/1511.03677

Lundberg, S. M., Nair, B., Vavilala, M. S., Horibe, M., Eisses, M. J., Adams, T., Liston, D. E., Low, D. K.-W., Newman, S.-F., Kim, J., & Lee, S.-I. (2018). Explainable machine-learning predictions for the prevention of hypoxaemia during surgery. Nature Biomedical Engineering, 2(10), 749–760. https://doi.org/10.1038/s41551-018-0304-0

Majumdar, S. R., Eurich, D. T., Gamble, J.-M., Senthilselvan, A., & Marrie, T. J. (2011). Oxygen saturations less than 92% are associated with major adverse events in outpatients with pneumonia: A population-based cohort study. Clinical Infectious Diseases: An Official Publication of the Infectious Diseases Society of America, 52(3), 325–331. https://doi.org/10.1093/cid/ciq076

McKinney, W. (2010). Data Structures for Statistical Computing in Python. Proceedings of the 9th Python in Science Conference, 56–61. https://doi.org/10.25080/Majora-92bf1922-00a

Nguyen, H., Jang, S., Ivanov, R., Bonafide, C. P., Weimer, J., & Lee, I. (2018). Reducing Pulse Oximetry False Alarms Without Missing Life-Threatening Events. Smart Health (Amsterdam, Netherlands), 9–10, 287–296. https://doi.org/10.1016/j.smhl.2018.07.002

Pedregosa, F., Varoquaux, G., Gramfort, A., Michel, V., Thirion, B., Grisel, O., Blondel, M., Prettenhofer, P., Weiss, R., Dubourg, V., Vanderplas, J., Passos, A., Cournapeau, D., Brucher, M., Perrot, M., & Duchesnay, É. (2011). Scikit-learn: Machine Learning in Python. Journal of Machine Learning Research, 12(85), 2825–2830.

Pollard, T. J., Johnson, A. E. W., Raffa, J. D., Celi, L. A., Mark, R. G., & Badawi, O. (2018). The eICU Collaborative Research Database, a freely available multi-center database for critical care research. Scientific Data, 5(1), 180178. https://doi.org/10.1038/sdata.2018.178

Sendelbach, S., & Funk, M. (2013). Alarm fatigue: A patient safety concern. AACN Advanced Critical Care, 24(4), 378–386; quiz 387–388. https://doi.org/10.1097/NCI.0b013e3182a903f9

Shenoy, N., Luchtel, R., & Gulani, P. (2020). Considerations for target oxygen saturation in COVID-19 patients: Are we under-shooting? BMC Medicine, 18(1), 260. https://doi.org/10.1186/s12916-020-01735-2

Sjoding, M. W., Dickson, R. P., Iwashyna, T. J., Gay, S. E., & Valley, T. S. (2020). Racial Bias in Pulse Oximetry Measurement. New England Journal of Medicine, 383(25), 2477–2478. https://doi.org/10.1056/NEJMc2029240

SRLF Trial Group. (2018). Hypoxemia in the ICU: Prevalence, treatment, and outcome. Annals of Intensive Care, 8. https://doi.org/10.1186/s13613-018-0424-4

Starr, N., Rebollo, D., Asemu, Y. M., Akalu, L., Mohammed, H. A., Menchamo, M. W., Melese, E., Bitew, S., Wilson, I., Tadesse, M., & Weiser, T. G. (2020). Pulse oximetry in low-resource settings during the COVID-19 pandemic. The Lancet Global Health, 8(9), e1121–e1122. https://doi.org/10.1016/S2214-109X(20)30287-4

Strachan, L., & Noble, D. W. (2001). Hypoxia and surgical patients—Prevention and treatment of an unnecessary cause of morbidity and mortality. Journal of the Royal College of Surgeons of Edinburgh, 46(5), 297–302.

Winters, B. D., Cvach, M. M., Bonafide, C. P., Hu, X., Konkani, A., O’Connor, M. F., Rothschild, J. M., Selby, N. M., Pelter, M. M., McLean, B., Kane-Gill, S. L., & Society for Critical Care Medicine Alarm and Alert Fatigue Task Force. (2018). Technological Distractions (Part 2): A Summary of Approaches to Manage Clinical Alarms With Intent to Reduce Alarm Fatigue. Critical Care Medicine, 46(1), 130–137. https://doi.org/10.1097/CCM.0000000000002803

World Health Organization. (2011). Pulse Oximetry Training Manual.

